# Factors associated with influenza vaccination among urban community-dwelling Chinese elderly: results from a multicity cross-sectional study

**DOI:** 10.1101/2025.08.12.25333514

**Authors:** Jiayue Guo, Xitong Jiao, Shuai Yuan, Lili You

## Abstract

**Background:** Influenza vaccination reduces morbidity and mortality in older adults. This study identifies characteristics and reasons for vaccination uptake among the elderly to inform strategies to improve coverage. Methods: We conducted a cross-sectional survey in December 2024 among community-dwelling adults aged ≥60 years across six Chinese cit-ies. Data collected included socio-demographic and health characteristics, influenza vac-cine awareness and uptake, reasons for vaccination or non-vaccination, and intentions for future vaccination. Univariate and multivariable logistic regression was used to identify factors associated with vaccination. To explore motivation patterns, co-occurrence net-works of vaccination reasons were constructed, and k-medoids clustering was applied. Results: Among 13,363 adults aged ≥60 years, influenza vaccination coverage was 34.0%. Higher education and income, being married, having health insurance, poor self-care ability, and chronic obstructive pulmonary disease were independently associated with vaccination. Vaccinated individuals reported more positive attitudes and were mainly motivated by family and doctor recommendations as well as perceived vaccine effective-ness, with four motivation profiles: social recommendation, comprehensive confidence, clinician-guided, and self-reliant confidence. Among unvaccinated participants, the main reasons for non-vaccination were mild influenza symptoms and the influence of family and friends, forming four motivation profiles: safety concern, low-perceived risk, social in-fluence, and perceived ineffectiveness. Conclusions: Influenza vaccination among older Chinese adults remains suboptimal. Tailored interventions leveraging healthcare provider endorsement, family and social support, and policy-driven strategies such as free or subsidized vaccination are needed, particularly for high-risk populations.

## 1. Introduction

Influenza is a highly contagious yet preventable viral respiratory illness that poses a substantial global health burden, causing considerable annual morbidity and mortality worldwide [1].Recent estimates suggest that approximately one billion seasonal influenza cases occur globally each year, including 3–5 million severe cases, resulting in 290,000–650,000 influenza-associated respiratory deaths[2]. Hospitalizations and deaths are largely concentrated among adults aged ≥ 60 years, who are more vulnerable due to age-related immune decline and a higher prevalence of chronic comorbidities[3]. A China-based study covering the 2010–2011 to 2014–2015 influenza seasons estimated an average of 88,100 excess respiratory deaths annually, with older adults accounting for 80% of these fatalities[4].

Influenza vaccination is a cost-effective strategy for reducing hospitalization and mortality rates among adults aged 65 and above[5]. Routine immunization also decreases outpatient visits for influenza-like symptoms and reduces expenditures on both prescription and over-the-counter medications. Therefore, promoting vaccination among older adults with chronic conditions represents a critical public health initiative. Although both the World Health Organization (WHO) and China’s Technical Guidelines for Influenza Vaccination recommend prioritizing adults aged 60 and older for influenza immunization[6], vaccination coverage among Chinese older adults remains suboptimal at 10%-30%, falling substantially below desired targets[7–10].

In many countries, annual influenza vaccination programs have been successfully integrated into national immunization strategies[11, 12]. However, it is noteworthy that in China, the influenza vaccine has not yet been included in the national immunization program and remains self-paid, with costs ranging from 60 to 80 RMB (approximately 9–15 USD). Only a few major cities offer local government-subsidized vaccination initiatives[13–16]. This situation significantly limits vaccine accessibility, particularly among elderly populations with limited awareness of influenza and constrained financial resources.

This study aims to characterize the socio-demographic profile of older adults receiving influenza vaccination in China and to analyze factors associated with vaccination uptake or non-vaccination, thereby providing evidence for optimizing future influenza vaccination strategies.

## 2. Materials and Methods

### 2.1. Study Design and Participants

In December 2024, we conducted a cross-sectional study across six Chinese cities—Beijing (North), Hangzhou (East), Qingdao (East), Shenzhen (South), Chongqing (Southwest), and Chengdu (Southwest)—selected to ensure broad geographical representation. In each city, five to eight Community Health Service Centers (CHSCs), responsible for providing vaccination services to urban residents, were randomly selected from a comprehensive local list using simple random sampling. Each CHSC invited 300 eligible individuals. Elderly residents aged ≥60 years attending CHSCs for free physical examinations or vaccinations were consecutively approached in waiting areas and invited to participate. Trained interviewers administered all questionnaires face-to-face using a digital system configured to require completion of all items before submission. Participants provided digital informed consent on the first page of the questionnaire. Each completed questionnaire was assigned a unique reference number and recorded in the system. In total, 13,754 individuals completed the questionnaire.

During the data processing phase, we applied strict inclusion criteria and logical consistency checks to screen the collected questionnaires. Invalid samples—such as those from respondents who did not meet the age threshold or provided contradictory responses—were removed to ensure the validity of the dataset. A total of 13,363 valid questionnaires were ultimately included, resulting in a valid response rate of 97.2%. The flow of study design and participant recruitment can be seen in Figure 1.

**Figure 1.**
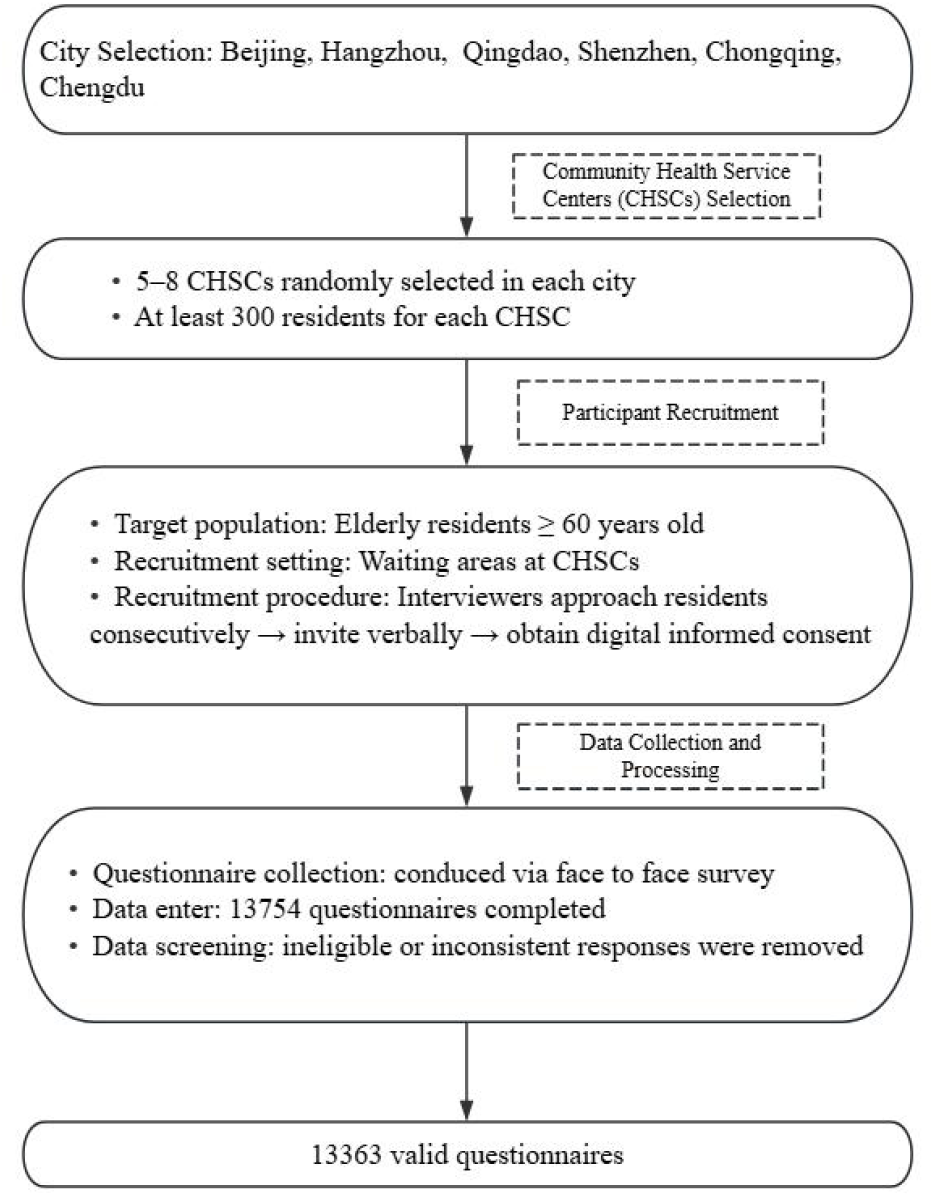
Flowchart of study design and participant recruitment.

### 2.2. Measures

The questionnaire comprised five sections: (i) socioeconomic and demographic variables; (ii) health status variables; (iii) awareness and attitudes toward influenza vaccination; (iv) reasons for accepting or declining vaccination; and (v) intention for revaccination. Details of the questionnaire are provided in Supplementary Material (File S1).

The outcome variable was prior influenza vaccination, assessed by the question, “Have you received an influenza vaccine in the past 12 months?” Responses were binary, with the response categories as “Yes” or “No”. Self-reported vaccination has been shown to be a reliable method for evaluating previous vaccination behavior among older adults [17].

Socio-demographic variables included region, gender, age, education, personal income, marital status, and health insurance coverage. Health status variables covered self-care ability, self-rated health status, number and type of chronic diseases, and self-reported indications for immunization. Attitudes were evaluated using 5-point Likert scales. Vaccinated respondents rated the perceived benefit of the vaccine from “very beneficial” to “not beneficial at all,” while unvaccinated respondents rated their perceived need for vaccination from “very necessary” to “Not necessary at all.” Multi-select questions were used to further explore specific reasons for vaccination or non-vaccination.

### 2.3. Statistically analysis

Descriptive analyses were conducted to provide an overview of socio-demographic and health-related variables. Influenza awareness and vaccination rates were calculated across different socio-demographic and health status subgroups. Categorical variables were compared using the χ^2^ test, while differences in attitudes between vaccinated and unvaccinated participants were assessed using t-tests.

To identify independent factors associated with influenza vaccination, univariate logistic regression analyses were performed, with socio-demographic and health-related characteristics as independent variables. Variables that showed statistical significance or marginal significance (p < 0.25, according to the Hosmer and Lemeshow criterion) in the univariate analysis were included in a multivariate logistic regression model using the Enter method. Results were presented as adjusted odds ratios (aORs) with 95% confidence intervals (CIs), and a reference category was specified for each variable. An aOR > 1 indicated a factor that facilitated vaccination, while an aOR < 1 indicated an inhibiting factor.

To explore co-occurrence patterns among vaccination reasons, separate networks were constructed for vaccinated and unvaccinated participants. Each network was represented as a binary matrix, with rows corresponding to participants and columns to specific reasons (coded as 1 if selected, 0 otherwise). Node-level centrality measures—including degree, betweenness, closeness, and eigenvector centrality—were calculated. Networks were visualized to display relationships among reasons and identify core factors.

To further analyze vaccination motivation patterns, reasons for both vaccinated and unvaccinated participants were examined using k-medoids clustering via the CLARA (Clustering Large Applications) algorithm. Jaccard distance was used to measure similarity between participants’ responses. Based on statistical considerations and interpretability, the number of clusters was pre-specified as four (k = 4). Each participant was assigned to a single cluster, representing a group with similar patterns of reasons for accepting or declining vaccination. Cluster-specific proportions of reason endorsement were then calculated and visualized in clustered heatmaps to identify distinct motivational profiles or reasons for non-vaccination.

All statistical analyses were conducted using R version 4.4 (R Foundation for Statistical Computing, Vienna, Austria) and IBM SPSS Statistics version 28 (IBM Corp., Armonk, NY, USA). Two-sided p-values < 0.05 were considered statistically significant.

## 3. Results

### 3.1. Subject Characteristics

The mean age of respondents was 69.53 years, with 45.3% male and 54.7% female. Participants were distributed across Chongqing (18.9%), Shenzhen (16.9%), Hangzhou (16.5%), Beijing (16.3%), Qingdao (16.0%), and Chengdu (15.4%). Overall, 66.6% of participants reported at least one chronic condition, with hypertension, diabetes, hyperlipidemia, and stroke being the most common comorbidities (see Table 1 for further sample characteristic). Influenza vaccine awareness was significantly associated with region, age, marital status, education level, income, self-care ability, self-rated health status, and the presence of hypertension or hyperlipidemia. In contrast, gender and certain chronic conditions—including diabetes, stroke, and chronic obstructive pulmonary disease (COPD)—were not significantly associated with vaccine awareness (Table 1).

**Table 1.**
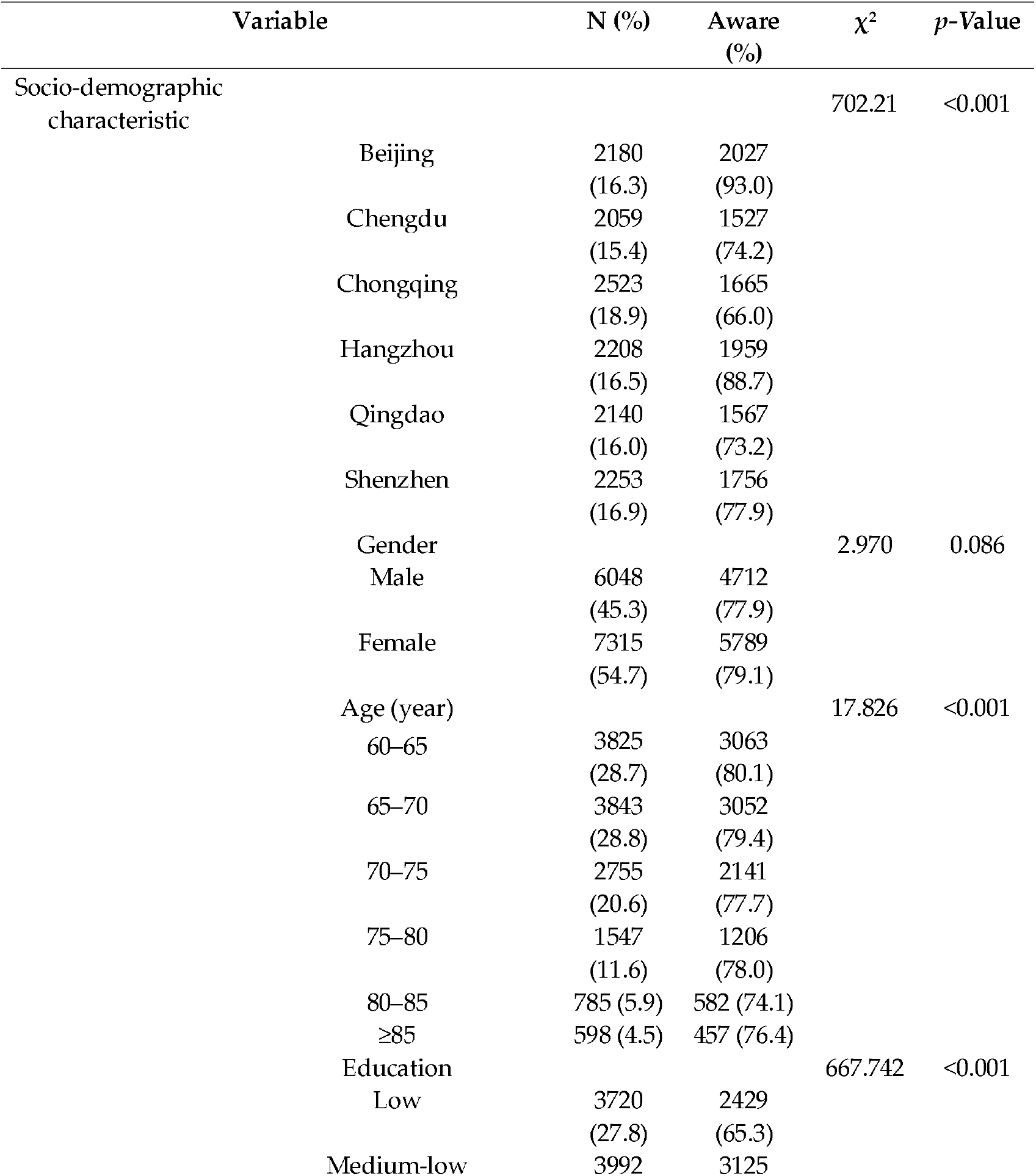
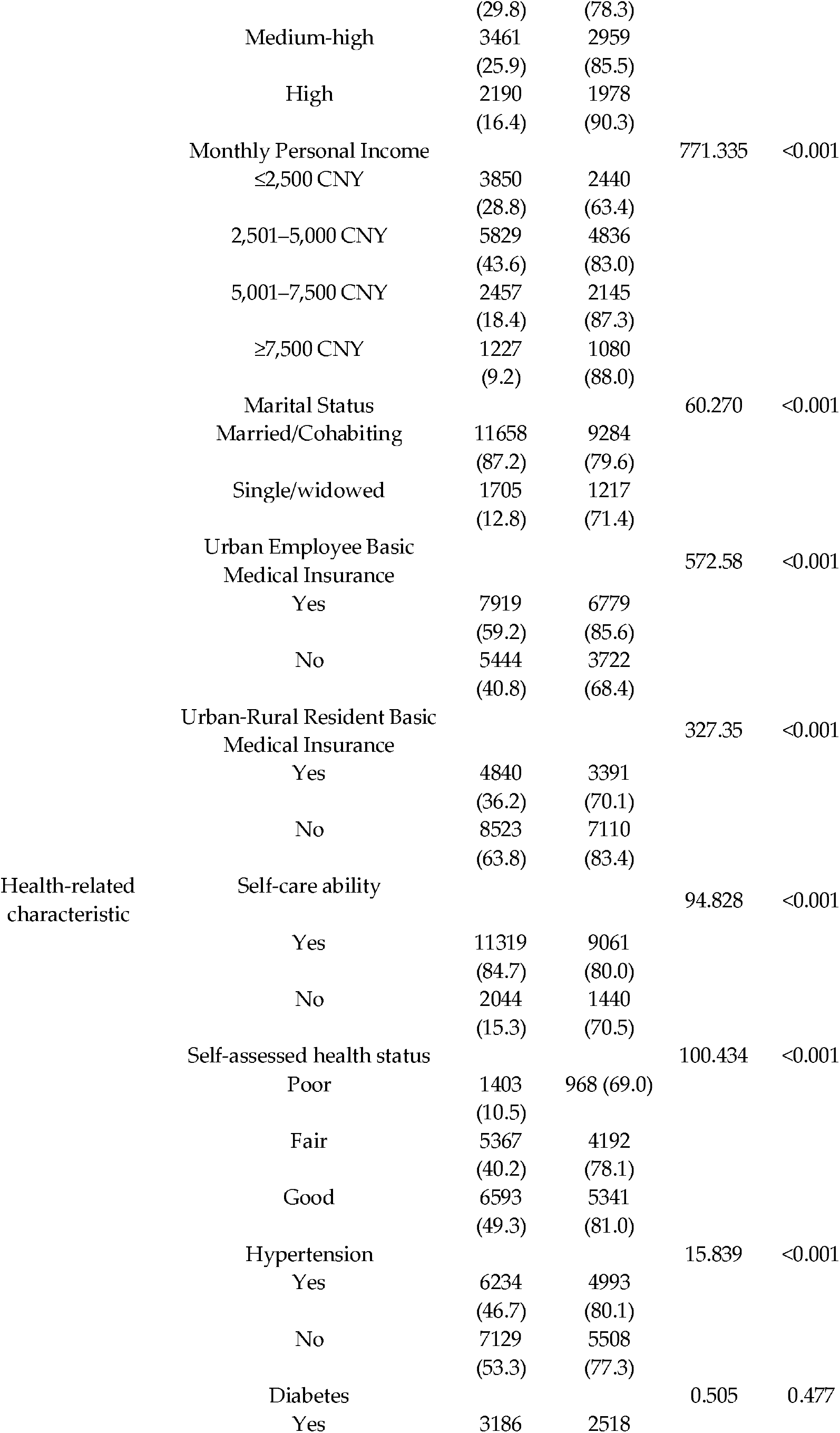
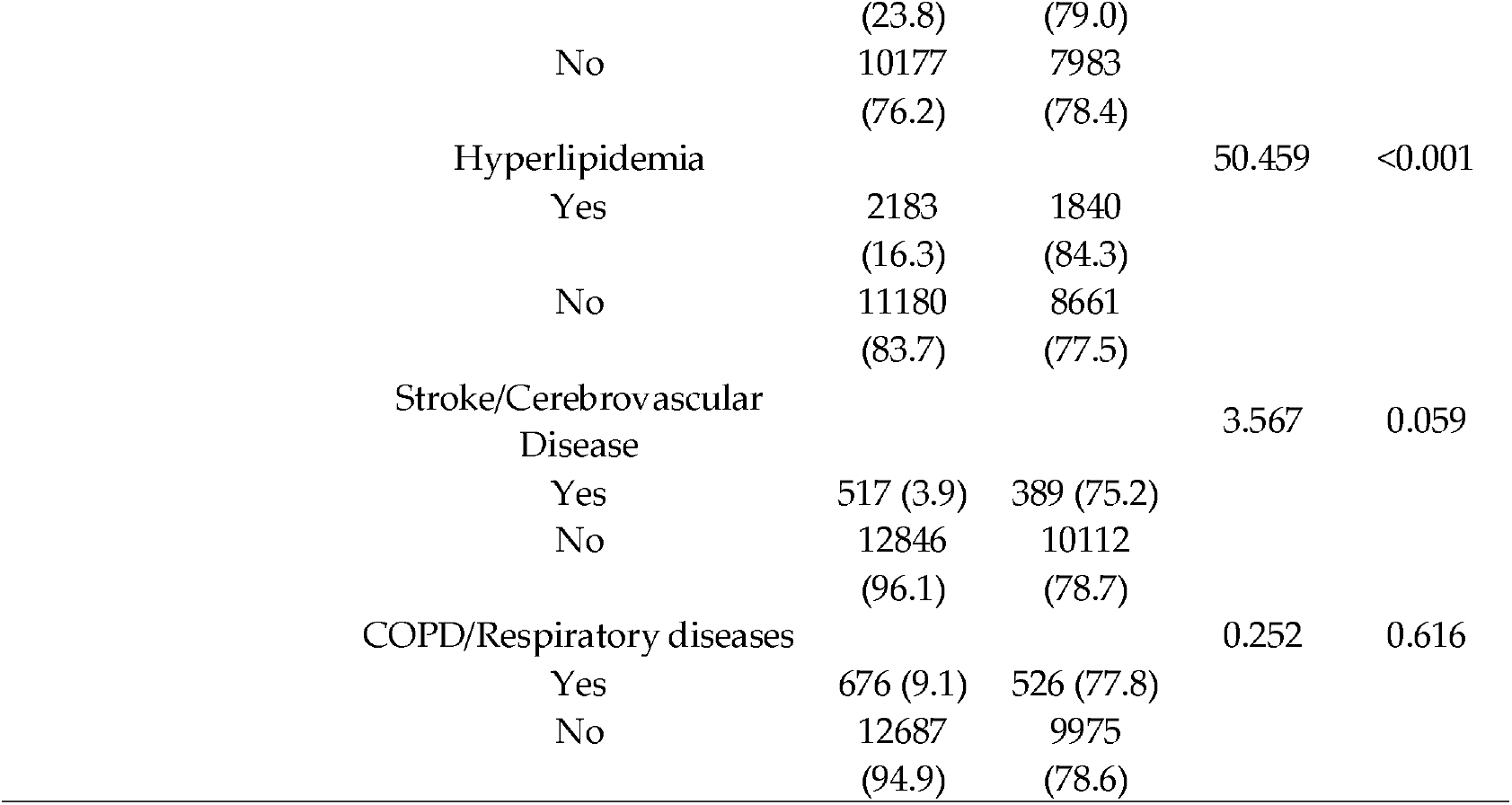
Influenza vaccine awareness by socio-demographic and health-related characteristics among urban community-dwelling Chinese adults aged ≥60 years (n = 13,363).

### 3.2. Vaccination Uptake

Overall, 34.0% of respondents reported having received an influenza vaccine within the past year. Among vaccinated individuals, 75.8% expressed willingness to be revaccinated the following year, whereas only 9.8% of unvaccinated participants intended to receive the vaccine.

Table 2 presents influenza vaccination rates stratified by socio-demographic and health-related characteristics. Among respondents aware of the influenza vaccine, 40.3% were vaccinated, compared with 11.1% among those unaware. Vaccination rates varied significantly across regions (p<0.001), with the highest rate observed in Hangzhou (45.9%), followed by Shenzhen (34.8%), Beijing (32.5%), Chongqing (31.7%), Chengdu (31.3%), and Qingdao (28.0%). No significant gender differences were observed (*p*=0.064), although males had a slightly higher vaccination rate than females (34.9% vs. 33.4%).

**Table 2.** Influenza vaccination rates by socio-demographic and health-related characteristics among urban community-dwelling Chinese adults aged ≥60 years (n = 13,363).

| Variable | Vaccinated N (%) | $\chi^2$ | p-Value |
| --- | --- | --- | --- |
| Awareness |  | 855.961 | <0.001 |
| Yes | 4233 (40.3) |  |  |
| No | 317 (11.1) |  |  |
| Region |  | 187.684 | <0.001 |
| Beijing | 708 (32.5) |  |  |
| Chengdu | 645 (31.3) |  |  |
| Chongqing | 801 (31.7) |  |  |
| Hangzhou | 1013 (45.9) |  |  |
| Qingdao | 600 (28.0) |  |  |
| Shenzhen | 783 (34.8) |  |  |
| Gender |  | 3.458 | 0.064 |
| Male | 2110 (34.9) |  |  |
| Female | 2440 (33.4) |  |  |
| Age (year) |  | 7.226 | 0.204 |
| 60–65 | 1313 (34.2) |  |  |
| 65–70 | 1327 (34.5) |  |  |
| 70–75 | 911 (33.1) |  |  |
| 75–80 | 545 (35.2) |  |  |
| 80–85 | 251 (32.0) |  |  |
| ≥85 | 44 (42.3) |  |  |
| Education |  | 98.662 | <0.001 |
| Low | 1043 (28.0) |  |  |
| Medium-low | 1383 (34.6) |  |  |
| Medium-high | 1256 (36.3) |  |  |
| High | 868 (39.6) |  |  |
| Monthly Personal Income |  | 157.152 | <0.001 |
| ≤2,500 CNY | 1001 (26.0) |  |  |
| 2,501–5,000 CNY | 2153 (36.9) |  |  |
| 5,001–7,500 CNY | 937 (38.1) |  |  |
| ≥7,500 CNY | 459 (37.4) |  |  |
| Marital Status |  | 14.897 | <0.001 |
| Married/Cohabiting | 4040 (34.7) |  |  |
| Single/widowed | 510 (29.9) |  |  |
| Urban Employee Basic Medical Insurance |  | 32.869 | <0.001 |
| Yes | 2850 (36.0) |  |  |
| No | 1700 (31.2) |  |  |
| Urban-Rural Resident Basic Medical Insurance |  | 11.167 | <0.001 |
| Yes | 1560 (35.1) |  |  |
| No | 2990 (32.2) |  |  |
| Self-care ability |  | 4.544 | 0.033 |
| Yes | 3812 (33.7) |  |  |
| No | 738 (36.1) |  |  |
| Self-assessed health status |  | 6.957 | 0.031 |
| Poor | 465 (33.1) |  |  |
| Fair | 1768 (32.9) |  |  |
| Good | 2317 (35.1) |  |  |
| Number of comorbidities |  | 12.040 | 0.007 |
| zero | 1470 (33.0) |  |  |
| One | 1721 (33.7) |  |  |
| Two | 833 (34.7) |  |  |
| Three or more | 526 (37.9) |  |  |
| Hypertension |  | 2.879 | 0.090 |
| Yes | 2169 (33.8) |  |  |
| No | 2381 (33.4) |  |  |
| Diabetes |  | 3.424 | 0.064 |
| Yes | 1128 (35.4) |  |  |
| No | 3422 (33.6) |  |  |
| Hyperlipidemia |  | 6.518 | 0.011 |
| Yes | 795 (36.4) |  |  |
| No | 3755 (33.6) |  |  |
| Stroke/Cerebrovascular Disease |  | 2.025 | 0.155 |
| Yes | 517 (31.1) |  |  |
| No | 12846 (34.2) |  |  |
| COPD/Respiratory diseases |  | 7.477 | 0.007 |
| Yes | 263 (38.9) |  |  |
| No | 4287 (33.8) |  |  |
| Self-reported Indicative of flu immunization |  | 2033.006 | 0.000 |
| Eligible | 3377 (74.2) |  |  |
| Ineligible | 1173 (25.8) |  |  |

Vaccination rates by age group were 34.2% for 60–64 years, 34.5% for 65–69 years, 33.1% for 70–74 years, 35.2% for 75–79 years, 32.0% for 80–84 years, and 42.3% for those aged ≥85 years, with no statistically significant differences between groups (*p*=0.204). Rates increased with education level (*p*<0.001), with the lowest rate among participants with primary education or less (28.0%), and higher rates among those with medium-low (34.6%), medium-high (36.3%), and high education levels (39.6%). Influenza vaccination rates were also positively associated with personal income (*p*<0.001).

Marital status was significantly associated with vaccination (*p*<0.001), with married or cohabiting adults showing a higher rate (34.7%) compared with single or widowed participants (29.9%). Vaccination rates were higher among holders of urban employee basic medical insurance (36.0%) and urban-rural resident basic medical insurance (35.1%) than among those without insurance.

Participants with chronic conditions generally had higher vaccination rates. Those with hypertension and diabetes had rates of 33.8% and 35.4%, respectively. Significant differences were observed for participants with hyperlipidemia (36.4%) and respiratory diseases, including COPD (38.9%). Vaccination rates also increased with the number of chronic conditions: 33.7% among those with one condition, 34.7% with two, and 37.9% with three or more.

Participants reporting better self-care ability had a slightly lower vaccination rate (*p*=0.033). Vaccination also varied by self-rated health status (*p*=0.031), with rates of 35.1%, 32.9%, and 33.1% among participants reporting “good,” “fair,” and “poor” health, respectively. Consistent with expectations, older adults meeting criteria for immunization demonstrated significantly higher vaccination rates.

### 3.3. Determinants of Vaccination

Regarding the socio-demographic factors affecting influenza vaccination among Chinese adults aged 60 years or older, individuals with medium-low education had 18.1% higher odds of vaccination (95% CI: 1.062-1.313, *p*=0.002), those with medium-high education had 19.3% higher odds (95% CI: 1.060-1.344, *p*=0.004), and those with high education had 38.2% higher odds (95% CI: 1.196-1.597, *p*<0.001), compared to those with low education. Higher monthly personal income was also associated with increased vaccination: compared with the ≤2,500 CNY group, individuals with an income of 2,501–5,000 CNY had 58.9% higher odds (95% CI: 1.432–1.764, *p*<0.001), those with 5,001–7,500 CNY had 58.3% higher odds (95% CI: 1.383–1.813, *p*<0.001), and those earning >7,500 CNY had 46.8% higher odds (95% CI: 1.236–1.743, *p*<0.001). Married/cohabiting individuals showed 1.15 times higher vaccination odds than single/widowed persons (95% CI: 1.026-1.296, *p*=0.016). Regarding health insurance, the OR for vaccination was 1.27 times higher (95% CI: 1.111-1.452, *p*<0.001) for those with Urban-Rural Resident Basic Medical Insurance, while Urban Employee Basic Medical Insurance showed no significant association with vaccination rate (95% CI: 0.983-1.287, *p*=0.088). No significant associations were observed between influenza vaccination rates and either gender or age after multivariate adjustment (Table 3).

**Table 3.** Socio-demographic factors affecting influenza vaccine uptake in the previous 12 months among urban community-dwelling Chinese adults aged ≥60 years (n=13363).

| Variables | Count | Univariate logistic regression | Multivariate logistic regression |
| --- | --- | --- | --- |

|  |  | OR | 95% CI | p-Value | OR | 95% CI | p-Value |
| --- | --- | --- | --- | --- | --- | --- | --- |
| Gender |  |  |  |  |  |  |  |
| Female | 7315 | 1 | 1 |  | 1 | 1 |  |
| Male | 6048 | 1.071 | 0.996-1.150 | 0.063 | 1.029 | 0.956-1.107 | 0.445 |
| Age |  |  |  |  |  |  |  |
| 60-<65 | 3835 | 1 | 1 |  | 1 | 1 |  |
| 65-<70 | 3843 | 1.013 | 0.922-1.113 | 0.787 | 1.107 | 0.925-1.118 | 0.729 |
| 60-<75 | 2755 | 0.949 | 0.855-1.053 | 0.322 | 0.982 | 0.884-1.091 | 0.737 |
| 75-<80 | 1547 | 1.045 | 0.923-1.182 | 0.388 | 1.123 | 0.990-1.275 | 0.071 |
| 80-<85 | 785 | 0.903 | 0.766-1.064 | 0.222 | 0.990 | 0.837-1.171 | 0.908 |
| ≥85 | 104 | 1.409 | 0.949-2.090 | 0.09 | 1.114 | 0.920-1.348 | 0.269 |
| Education |  |  |  |  |  |  |  |
| Low | 3720 | 1 | 1 |  | 1 | 1 |  |
| Medium-low | 3992 | 1.361 | 1.235-1.499 | <0.001 | 1.181 | 1.062-1.313 | 0.002 |
| Medium-high | 3461 | 1.462 | 1.321-1.615 | <0.001 | 1.193 | 1.060-1.344 | 0.004 |
| High | 2190 | 1.462 | 1.507-1.884 | <0.001 | 1.382 | 1.196-1.597 | <0.001 |
| Monthly Personal Income |  |  |  |  |  |  |  |
| ≤2,500 CNY | 3850 | 1 | 1 |  | 1 | 1 |  |
| 2,501-5,000 CNY | 5825 | 1.667 | 1.524-1.823 | <0.001 | 1.589 | 1.432-1.764 | <0.001 |
| 5,001-7,500 CNY | 2457 | 1.755 | 1.574-1.956 | <0.001 | 1.583 | 1.383-1.813 | <0.001 |
| 7,500 CNY | 1227 | 1.701 | 1.484-1.949 | <0.001 | 1.468 | 1.236-1.743 | <0.001 |
| Marital Status |  |  |  |  |  |  |  |
| Single/widowed | 1705 | 1 | 1 |  | 1 | 1 |  |
| Married/Cohabiting | 11658 | 1.243 | 1.113-1.388 | <0.001 | 1.153 | 1.026-1.296 | 0.016 |
| Urban Employee Basic Medical Insurance |  |  |  |  |  |  |  |
| No | 5446 | 1 | 1 |  | 1 | 1 |  |
| Yes | 7917 | 1.239 | 1.152-1.334 | <0.001 | 1.124 | 0.983-1.287 | 0.088 |
| Urban-Rural Resident Basic Medical Insurance |  |  |  |  |  |  |  |
| No | 8523 | 1 | 1 |  | 1 | 1 |  |
| Yes | 4840 | 1.136 | 1.054-1.225 | <0.001 | 1.270 | 1.111-1.452 | <0.001 |

Table 4 shows the health status factors that affect influenza vaccination among Chinese adults aged 60 years older. After multivariate adjustment, individuals with poor self-care ability had 15.4% higher vaccination odds compared to those with self-care ability (95% CI: 1.041-1.280, *p*=0.006). Regarding self-assessed health status, those reporting good health showed 21.8% higher vaccination odds than those with poor health (95% CI: 1.067-1.391, *p*=0.003). For chronic conditions, the OR was 1.13 times higher (CI: 1.026-1.251, *p*=0.014) in the presence of hyperlipidemia and 1.294 times higher (95% CI: 1.101-1.521, *p*=0.002) in the presence of respiratory diseases or COPD. However, no significant associations were found for hypertension, diabetes, and cerebrovascular disease or stroke after multivariate adjustment.

**Table 4.** Health status factors affecting influenza vaccine uptake in the previous 12 months among urban community-dwelling Chinese adults aged ≥60 years (n=13363).

| Risk factor | Coun | Univariate logistic | Multivariate logistic |
| --- | --- | --- | --- |

|  | t | OR | regression<br>95% CI | p-Value | OR | regression<br>95% CI | p-Value |
| --- | --- | --- | --- | --- | --- | --- | --- |
| Self-care ability |  |  |  |  |  |  |  |
| Yes | 11319 | 1 | 1 |  | 1 | 1 |  |
| No | 2044 | 1.11<br>3 | 1.009-1.22<br>8 | 0.033 | 1.15<br>4 | 1.041-1.28<br>0 | 0.006 |
| Self-assessed health status |  |  |  |  |  |  |  |
| Poor | 1403 | 1 | 1 |  | 1 | 1 |  |
| Average | 5367 | 0.99<br>1 | 0.875-1.12<br>3 | 0.887 | 1.04<br>4 | 0.917-1.18<br>7 | 0.517 |
| Good | 6593 | 1.09<br>3 | 0.967-1.23<br>5 | 0.153 | 1.21<br>8 | 0.067-1.39<br>1 | 0.003 |
| Hypertension |  |  |  |  |  |  |  |
| No | 7129 | 1 | 1 | 0.090 | 1 | 1 |  |
| Yes | 6234 | 1.06<br>4 | 0.990-1.14<br>3 |  | 1.06<br>8 | 0.991-1.15<br>2 | 0.084 |
| Diabetes |  |  |  |  |  |  |  |
| No | 10177 | 1 | 1 |  | 1 | 1 |  |
| Yes | 3186 | 1.08<br>2 | 0.995-1.17<br>6 | 0.064 | 1.08<br>3 | 0.993-1.18<br>2 | 0.073 |
| Hyperlipidemia |  |  |  |  |  |  |  |
| No | 11180 | 1 | 1 |  | 1 | 1 |  |
| Yes | 2183 | 1.13<br>3 | 1.029-1.24<br>6 | 0.010 | 1.13<br>3 | 1.026-1.25<br>1 | 0.014 |
| Stroke/Cerebrovascular Disease |  |  |  |  |  |  |  |
| No | 12846 | 1 | 1 |  | 1 | 1 |  |
| Yes | 517 | 0.87<br>1 | 0.721-1.05<br>3 | 0.155 | 0.84<br>8 | 0.697-1.03<br>1 | 0.098 |
| COPD/Respiratory diseases |  |  |  |  |  |  |  |
| No | 12687 | 1 | 1 |  | 1 | 1 |  |
| Yes | 676 | 1.24<br>8 | 1.064-1.463 | 0.006 | 1.29<br>4 | 1.101-1.52<br>1 | 0.002 |

### 3.4. Vaccination Motivations and Reasons

Vaccinated participants reported a significantly higher mean attitude score (mean = 4.03) compared with unvaccinated participants (mean = 2.61; *p* < 0.001), indicating more positive perceptions of vaccine benefits among those who had been vaccinated. See details in Supplementary Material (File S2).

Table 5 presents the node-level centrality measures of vaccination reasons in the co-occurrence network constructed from vaccinated participant responses. All reasons had a degree of 7, indicating frequent co-occurrence with other reasons.

**Table 5.** Network analysis of co-occurring reasons for vaccination among vaccinated participants.

| Reason | Degree | Betweenness | Closeness | Eigenvector |
| --- | --- | --- | --- | --- |
| Safe/effective | 7 | 0 | 8.278 | 0.954 |
| Family recommendation | 7 | 0 | 7.610 | 1.000 |
| Doctor recommendation | 7 | 0 | 8.756 | 0.820 |
| Social media | 7 | 14 | 0.002 | 0.283 |
| Prior helpful | 7 | 0 | 0.001 | 0.687 |
| Government recommendation | 7 | 0 | 9.930 | 0.530 |
| Protect family | 7 | 0 | 0.001 | 0.572 |
| Affordable | 7 | 0 | 0.001 | 0.330 |

“Family recommendation”, “Safe/effective”, and “Doctor recommendation” exhibited the highest eigenvector centrality, suggesting they are the core drivers of vaccination decisions. “Social media” showed a high betweenness centrality but lower eigenvector centrality, indicating its role as a bridge connecting different reasons rather than as a core factor. Other reasons, including “Government recommendation”, “Protect family”, and “Affordable”, were moderately connected, contributing to overall network structure but with less central influence. The structure and relationships among these reasons are visualized in the co-occurrence network diagram (Figure 2).

**Figure 2.**
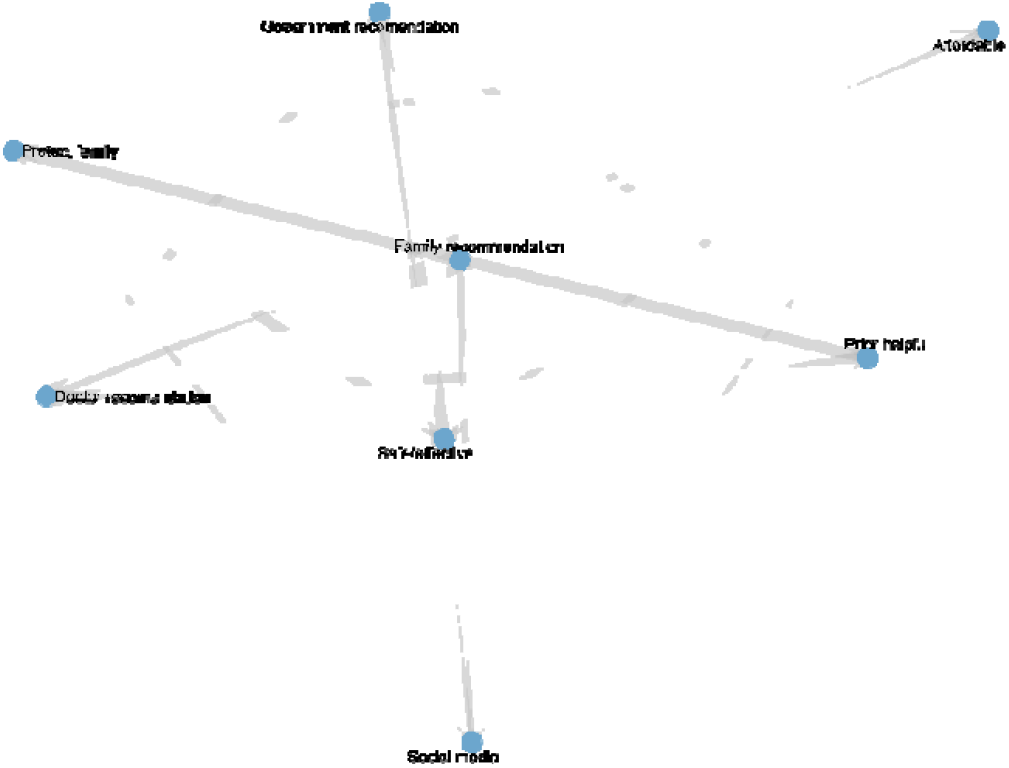
Network of vaccination reasons among vaccinated participants.

Table 6 shows node-level centrality measures of non-vaccination reasons in the co-occurrence network among unvaccinated participants. All reasons had a degree of 8, indicating frequent co-selection. Core reasons included “Mild influenza symptoms”, “Concerns about side effects”, “Perceived vaccine ineffectiveness”, and “Family and friends influence”, reflecting the main barriers to vaccination. “Doctor recommendation” had the highest betweenness centrality but lower eigenvector centrality, acting as a bridge connecting different combinations of reasons. Other reasons, such as “Social media influence”, “Commercial motives”, “Access inconvenience”, and “High vaccination cost”, were moderately connected, contributing to the network structure but with less central influence. The detailed structure and relationships of these vaccination reasons are illustrated in the co-occurrence network diagram (Figure 3).

**Table 6.**
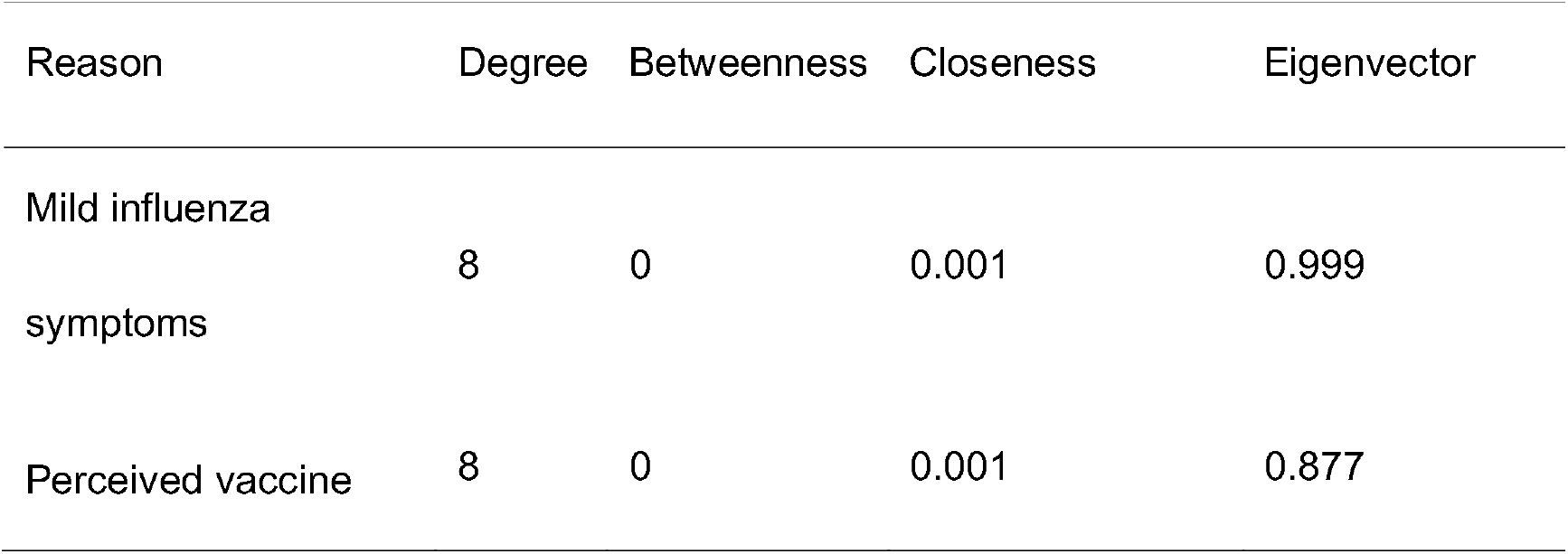
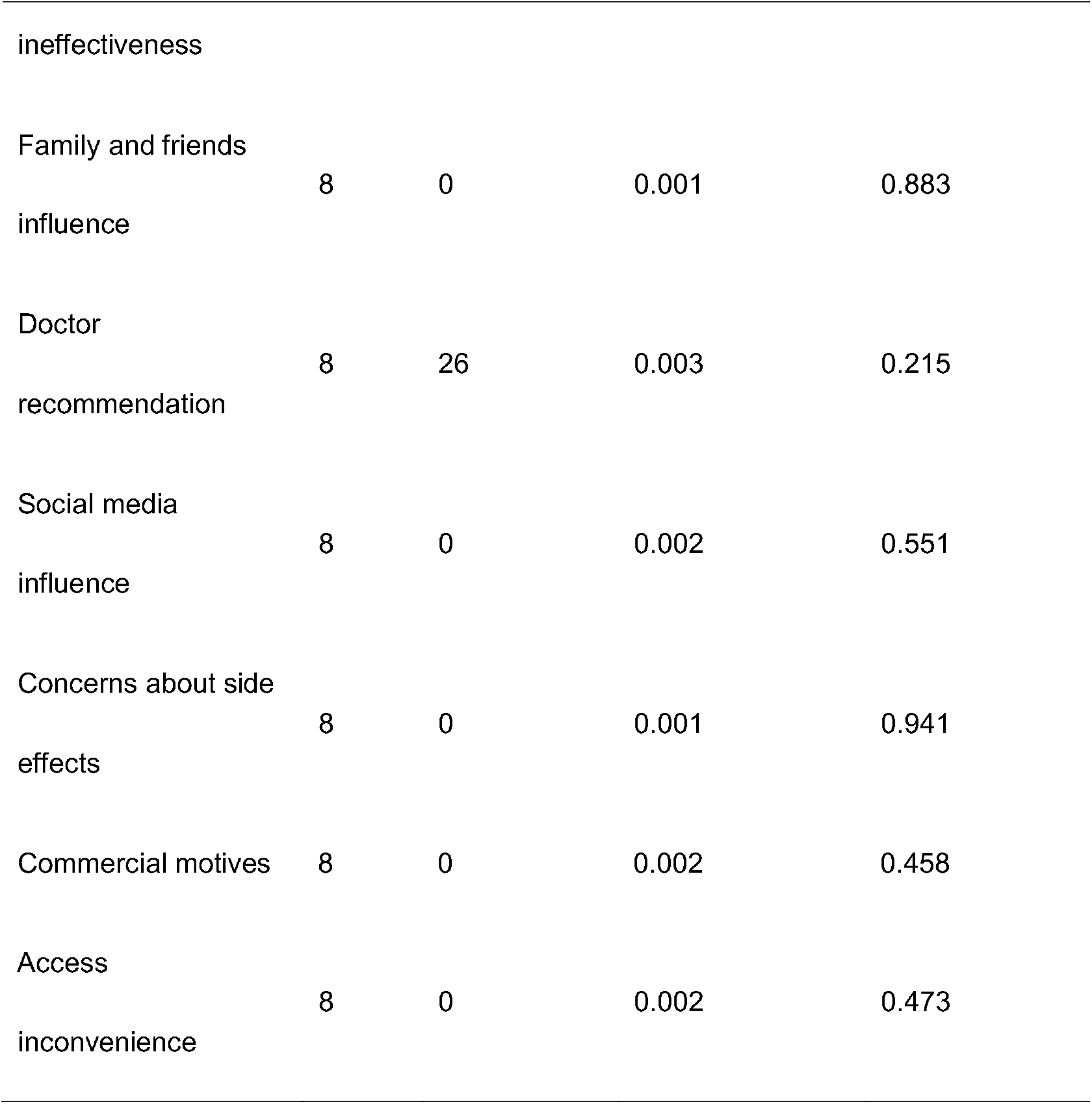
Network analysis of co-occurring reasons for non-vaccination among unvaccinated participants.

**Figure 3.**
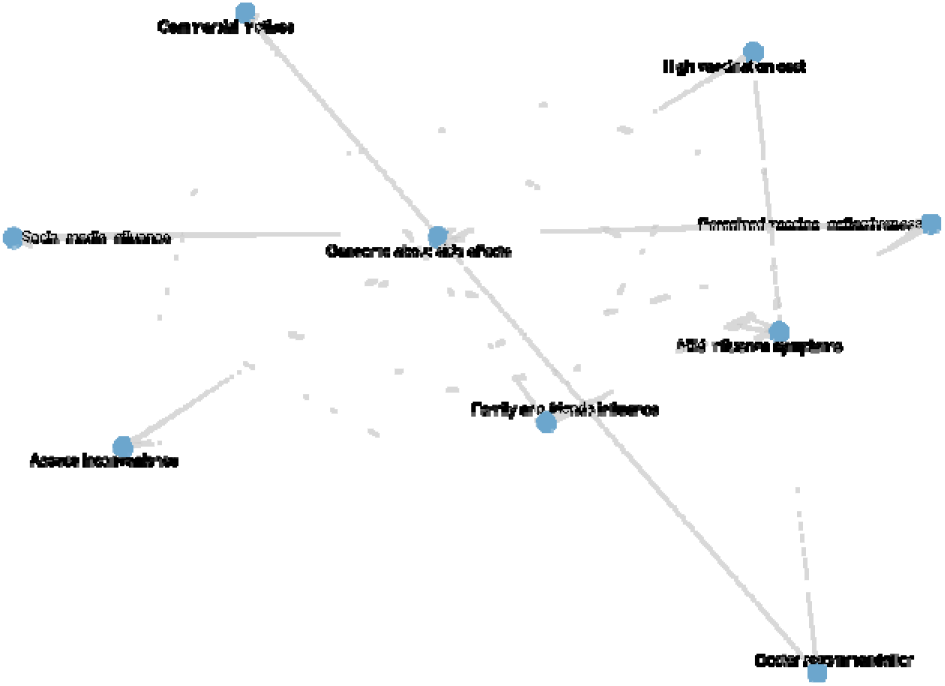
Network of reasons for non-vaccination among unvaccinated participants.

Among vaccinated participants, the most frequently selected reason for vaccination was “safe and effective” (n=2222, 48.8%), followed by “family recommendation” (n=1617, 35.5%) and “doctor recommendation” (n=1125, 24.7%). Other reported reasons included “prior helpful experience” (n=790, 17.4%), “government recommendation” (n=522, 11.5%), “protecting family” (n=484, 10.6%), “social media” (n=282, 6.2%), and “affordable” (n=241, 5.3%). Vaccination reasons were further analyzed using k-medoids clustering based on Jaccard distance, identifying four clusters (k = 4), each representing a distinct motivational profile (Supplementary Material, File S3). The distribution of reasons across clusters is visualized in a clustered heatmap (Figure 4).

**Figure 4.**
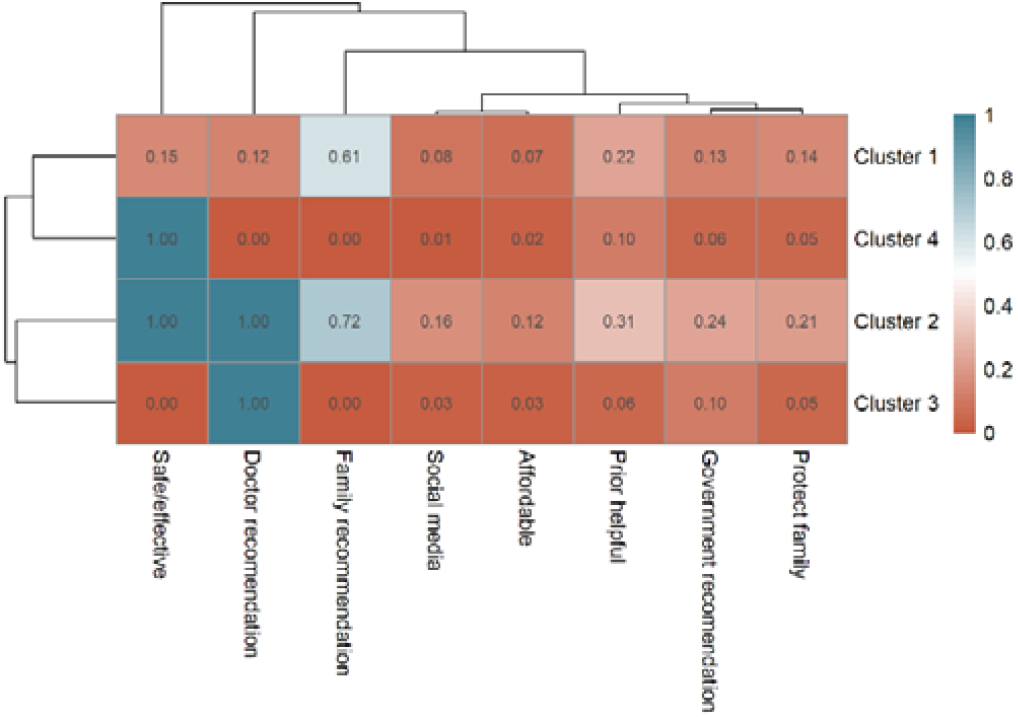
Clustered heatmap of vaccination motivations among vaccinated participants showing four distinct motivation profiles.

Cluster 1 (Family & Social Recommendation Group, n = 2199) represented individuals primarily influenced by family recommendations (mean = 0.61), indicating reliance on social trust rather than professional guidance. Cluster 2 (Comprehensive Confidence Group, n = 393) exhibited the highest endorsement across almost all motivations, including perceived vaccine safety (1.00) and doctor recommendation (1.00), reflecting multi-faceted confidence. Cluster 3 (Clinician-Guided Group, n = 462) was predominantly driven by medical advice (doctor recommendation = 1.00), with minimal influence from other sources. Cluster 4 (Self-Reliant Confidence Group, n = 1496) was characterized by autonomous belief in vaccine effectiveness, reflecting rational self-decision.

Among unvaccinated participants, the most frequently selected reason for not receiving the vaccine was “mild influenza symptoms” (n=3,544, 40.2%), followed by “family and friends influence” (n=2,079, 23.6%), “concerns about side effects” (n=1,642, 18.6%), and “perceived vaccine ineffectiveness” (n=1,526, 17.3%). Other reported reasons included “high vaccination cost” (n=811, 9.2%), “access inconvenience” (n=731, 8.3%), “social media influence” (n=590, 6.7%), “doctor recommendation” (n=573, 6.5%), and “commercial motives” (n=520, 5.9%). Clustering analysis similarly identified four distinct participant groups (Supplementary Material, File S4), visualized in a clustered heatmap (Figure 5):

**Figure 5.**
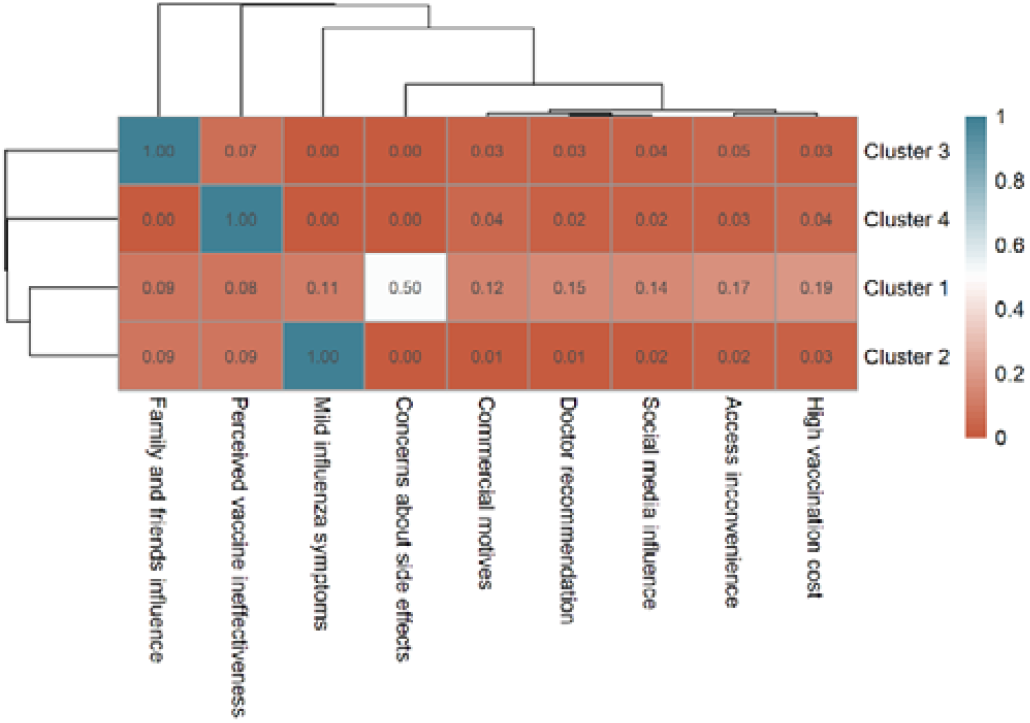
Clustered heatmap of non-vaccination motivations among unvaccinated participants showing four distinct motivation profiles.

Cluster 1 (Safety Concern Group, n = 3,258) showed a high frequency for concerns about side effects (mean = 0.50), representing participants mainly deterred by safety-related issues. Cluster 2 (Low-Perceived Risk Group, n = 3,189) was characterized by mild influenza symptoms (mean = 1.00), suggesting participants who did not perceive influenza as a serious illness. Cluster 3 (Social Influence Group, n = 1,518) had the highest frequency for family and friends influence (mean = 1.00), indicating decisions primarily shaped by social opinions or discouragement from peers. Cluster 4 (Perceived Ineffectiveness Group, n = 848) reported perceived vaccine ineffectiveness as the main reason for not receiving vaccination.

## 4. Discussion

This multicity cross-sectional study investigated influenza vaccination uptake in the past year among community-dwelling older adults in urban China and identified heterogeneous profiles of vaccination motivations and barriers.

This study reveals that influenza vaccination coverage among urban Chinese adults aged 60 and above remains suboptimal (28.0%-45.9%), falling substantially below the World Health Organization’s 75% target and lower than vaccination rates observed in developed countries [18, 19]. Previous research has also shown generally low vaccination coverage among older populations in China[15]. A systematic review reported an average vaccination rate of 51.29% among adults aged 60 and above, with Belgium ranking highest at 72.73%, followed by Hong Kong SAR, China, at 54%[20]. These findings underscore the urgent need to identify factors associated with vaccination behavior and implement community-based policy promotion to improve immunization uptake.

China has implemented heterogeneous reimbursement policies for influenza vaccination across different regions. In most areas, vaccination remains self-paid. Since 2020, Hangzhou has provided free influenza vaccines to adults aged 70 and above. This study demonstrates that Hangzhou achieved significantly higher vaccination coverage compared to other cities, indicating that policy-driven financial accessibility serves as a core driver for vaccine uptake[21]. Although the COVID-19 pandemic heightened public willingness for vaccination, only cost-free provision effectively translates this intention into actual vaccination behavior[22]. Governments or other service organizations should consider subsidized services and vigorously promote free influenza vaccination policies[10].

This study demonstrates that educational attainment, income/socioeconomic status, marital status, and health insurance coverage are significant determinants of influenza vaccination among older adults in China. Insured individuals showed higher vaccination rates, consistent with findings from North American studies[23]. Substantial evidence confirms the role of socio-demographic factors[24]: those cohabiting with spouses or family members exhibited higher vaccination rates[25, 26], as social networks and family recommendations positively influence health information acquisition[27]. Increased age was associated with increased vaccination uptake, while higher income[25] and elevated educational attainment[25, 28, 29] or socioeconomic status[30, 31] were identified as potential facilitators for vaccination among the elderly. Notably, we found no significant association between gender and vaccination rates, whereas international evidence remains divergent: multiple studies report higher coverage among males[32–36], others indicate female predominance[23, 37–39], and some show no significant gender differences[40].

Poorer health status—including limited self-care ability, multiple chronic diseases, hyperlipidemia, and respiratory conditions—increases vaccination uptake among older adults. These results are consistent with studies from Europe[41] and Singapore[42], where more chronic diseases correlated with higher vaccination rates. Multiple studies confirm that health conditions, especially specific chronic diseases[39, 43–47], influence vaccination in the elderly. A systematic review also found higher flu vaccination rates in chronic disease patients globally than in the general population[20]. Due to their greater vulnerability to flu and higher risk of severe complications—such as worsening of existing diseases, pneumonia, multi-organ failure, and death—vaccination is especially important for this group. COPD stood out as a key factor, likely because these patients have weaker cellular immune responses to flu and depend more on vaccine-protected antibody immunity. To enhance vaccination coverage, targeted strategies should focus on high-risk populations, including those with chronic conditions such as hyperlipidemia or COPD, as well as socioeconomically disadvantaged groups. As this study draws on representative samples from six major Chinese cities, its findings demonstrate robust credibility and reliability, providing valuable supplementary evidence to existing research.

Among vaccinated participants, four main motivation types were identified: family recommendation, comprehensive confidence, clinician-guided, and self-reliant confidence. family recommendation participants relied on family advice rather than professional guidance; comprehensive confidence participants showed high motivation across multiple factors; clinician-guided participants were primarily influenced by physicians; and self-reliant confidence participants trusted vaccine safety independently. Among unvaccinated participants, four barrier types emerged: Safety Concern, Low Risk Perception, Social Influence, and Perceived Vaccine Ineffectiveness. Safety concerns, low perceived risk, social influence, and doubts about efficacy were the main reasons for non-vaccination. Overall, vaccinated individuals were driven by external guidance, while unvaccinated individuals were influenced mainly by safety, risk perception, and social factors. These findings highlight the need to tailor vaccination strategies to the specific motivations and barriers of different groups.

Failure to recognize the increased susceptibility to influenza resulting from the combined effects of age and medical risk factors is a primary reason for reduced vaccination willingness. Low intention to vaccinate was also associated with insufficient awareness of the benefits of influenza vaccination and concerns about vaccine safety. Such health beliefs have indeed been identified in international studies as prominently affecting influenza vaccination behaviors among the general older population[48, 49]. The study also revealed that recommendations from healthcare providers and influence from family and friends are key factors promoting vaccination behavior. Social integration was found to positively contribute to vaccination uptake[50]. Therefore, the next critical step to improving vaccination rates lies in leveraging the endorsement and recommendations of medical professionals to address the two major barriers—misconceptions about influenza and concerns about vaccine safety—while also improving knowledge and attitudes. Emphasis should be placed on the supportive role of family and informal social networks. Primary healthcare institutions should not only ensure service accessibility but also allocate sufficient consultation time to thoroughly discuss the benefits and risks of vaccine protection with patients. Furthermore, regular health education initiatives on influenza and its vaccination should be conducted to enhance awareness, attitudes, and practices among older adults.

Consistent with previous research findings[51, 52], we find that prior influenza vaccination experience significantly promotes subsequent vaccination behaviour among older adults in China. A notable cognitive difference was also observed between vaccinated and unvaccinated groups: over 70% of vaccinated individuals affirmed the vaccine’s effectiveness and expressed willingness to be revaccinated the following year, whereas nearly 90% of unvaccinated older adults reported no intention to vaccinate in the coming year. These results suggest that health administrative departments should seize critical windows for intervention. Encouraging currently unvaccinated individuals to receive the vaccine may initiate a “carry-over effect”, promoting sustained vaccination in subsequent seasons.

The routine medical follow-ups received by older adults present a valuable opportunity for promoting vaccination. Organized immunization counseling in primary healthcare settings can provide evidence-based information to correct misconceptions, biases, and negative attitudes toward vaccination. Counseling should include objective facts such as that influenza vaccines are not 100% effective[53] and may have side effects[54]. Emphasis should be placed on educating high-risk groups about how influenza can exacerbate their underlying conditions, while annual vaccination can effectively reduce infection and prevent severe complications. Simple interventions such as personalized postcards or telephone reminders[55], as well as recommendations from healthcare professionals[56–61], can positively influence vaccination rates. Practice has demonstrated that home-visit vaccination services can also effectively improve coverage. Godoy et al. [62] specifically highlighted that patients whose physicians had been vaccinated showed significantly higher vaccination rates than those whose physicians had not. At the systems level, policy interventions may further enhance vaccination rates. Population-based influenza vaccination strategies can also leverage information systems to enable efficient tracking, vaccination, and monitoring of high-risk groups.

## 5. Conclusions

Our findings indicate that influenza vaccination coverage among older adults in China remains below the World Health Organization’s target. Higher uptake was observed in socioeconomically advantaged groups with better access to healthcare, often despite poorer functional health. Targeted interventions are needed for unmarried, less-educated, and uninsured older adults. Major barriers include misconceptions about influenza, social influence from unvaccinated peers or family, and concerns regarding vaccine safety and effectiveness. Strategies to improve coverage should leverage primary healthcare services, structured immunization counseling, family and social networks, personalized reminders, and policy-supported financial incentives.

## Supplementary Materials

File S1: Questionnaire Structure and Content; File S2: Attitude scores toward influenza vaccination among vaccinated and unvaccinated participants; File S3: Proportions of vaccination reasons endorsed within each cluster among vaccinated participants; File S4: Proportions of non-vaccination reasons endorsed within each cluster among unvaccinated participants.

## Author Contributions

Conceptualization, J.G. and L.Y.; Methodology, J.G.; formal Analysis, J.G.; Investigation, J.G., X.J and S.Y; Writing – Original Draft Preparation, J.G.; Writing – Review & Editing, J.G., X.J. and S.Y; Supervision, L.Y.; Funding Acquisition, L.Y. All writers affirm they satisfy the ICMJE criteria for authorship and made significant contributions to the conception and design, participated in writing the article or revising it critically for significant intellectual content, and gave final approval to its submission. All authors have read and agreed to the published version of the manuscript.

## Funding

This study was funded by the Beijing Natural Science Foundation-Haidian Original Innovation Joint Fund Project [grant number L242146].

## Institutional Review Board Statement

This study was conducted according to the guidelines laid down in the Declaration of Helsinki and obtained ethical approval from the Institutional Board of the Chinese Academy of Medical Sciences & Peking Union Medical College, which oversees research involving human subjects on 21 January 2025 (CAMS&PUMC-IEC-2025-002).

## Informed Consent Statement

Informed consent was obtained from all subjects involved in the study.

## Data Availability Statement

The data within this article will be shared upon reasonable request to the corresponding author.

## Acknowledgments

We are deeply appreciative of all the participants from Beijing, Shenzhen, Hangzhou, Qingdao, Chongqing, and Chengdu for their invaluable time and insights. We also extend our heartfelt gratitude to our postgraduate research assistants at Peking Union Medical College for their unwavering cooperation and assistance.

## Conflicts of Interest

The authors have no conflicts of interest relevant to this article to disclose.

## Notes

### Competing Interest Statement

The authors have declared no competing interest.

### Funding Statement

the Beijing Natural Science Foundation (BNSF)

### Author Declarations

the Institutional Review Board of the Chinese Academy of Medical Sciences & Peking Union Medical College

### Summary of Updates

This version of the manuscript has been revised to update the methodology and figures.

